# Human Metapneumovirus seroprevalence and PCR trends - Findings from a Tertiary Care Hospital in India

**DOI:** 10.1101/2025.02.12.25322117

**Authors:** Sadhana Yelamanchili, Deepika Gujjarlapudi, Ankit Mittal, Nagamani Chella, Sushma Yadav, Venkata Rajesh Dulla, Nageshwar reddy Duvvur

## Abstract

Human Metapneumovirus (hMPV) is a paramyxovirus that was detected in India first in 2004. It causes a mild common cold in most cases but serious illness has been reported in some young children, adults >65 years and the immunocompromised.

**AIM:** To study the age related prevalence of IgG antibodies in the population and also to determine the percent positivity and seasonal variation of hMPV infection.

**MATERIAL AND METHODS:** This cross-sectional study assessed seroprevalence by testing 820 serum samples collected between January 25^th^ – 31^st^ 2025 for hMPV IgG antibodies by ELISA and stratified according to age and sex. hMPV PCR trends: Retrospective data from the 1276 tests for Respiratory Viruses done at our centre during 2023 and 2024 were analysed and comprised of age groups 0-18, 19-30 ,31-45, 45-60 and > 60 years. The relationship between gender (M/F) and outcome (positive/negative) across various age groups was analysed using the chi-square test for independence in Graph Pad Prism 8.0.

**RESULTS: Seroprevalence study:** Overall hMPV IgG Antibody Positivity was 53.4%. Antibody positivity was higher in above 60 years when compared to other groups and was statistically significant (P=0.0001) and female predominance is seen.

**In hMPV PCR trends:** A higher percentage of positivity were observed in > 60years individuals who had hospitalisation when compared to the other groups which was statistically significant(P=0.0001). Overall Percent positivity was 7.8% in 2023 and 3.8% in 2024. Seasonal peaks occurred in Feb-Mar and Oct. 92.6% of patients were discharged and doing well on follow-up and only 7.4% deaths were seen.

**CONCLUSION:** Our findings highlight around 53.4% of the total study population had hMPV IgG antibodies. PCR positivity and antibody positivity was higher in individuals over 60 who had other comorbidities. Hospitalization and mortality rates are significantly high in this high risk groups. Vaccine development for high-risk individuals is recommended.

## INTRODUCTION

Human Metapneumovirus (hMPV) is one of the viruses that cause the common cold. Most people are infected by 5 years of age. Most cases are mild, but young children, adults over 65 and people with weakened immune systems are at a higher risk of serious illness.

The virus is an enveloped virus and classified in the Paramyxovirus family (pneumoviridae subfamily). It was first identified in 2001 in Netherlands. It was detected in India in 2004. It is found around the world. Two major strains, designated A and B each with two subtypes have been identified, by antigenic and genetic analysis. ^(1)^ N. Devenathan et al ^(9)^ in their study reported the circulation of emerging lineages A2.2.1& A2.2.2 among children under 5 years of age in south India.

Despite its global prevalence, the Indian population’s hMPV IgG seroprevalence data is sparse. Amid reports of increased hMPV cases in China, we investigated age-related hMPV IgG antibody prevalence, PCR trends and seasonal variations, aiming for respiratory outbreak preparedness and public health interventions.

## MATERIALS AND METHODS

### Study Design

This was a cross-sectional study conducted at AIG Hospitals, Hyderabad, India, a high-volume multispecialty tertiary care center that also served as a designated COVID-19 center during the pandemic. The study comprised two parts:

### hMPV IgG Seroprevalence Study

A total of 820 serum samples from the departments of Serology and Biochemistry, collected between January 25th and 31st, 2025, were tested for hMPV IgG antibodies using the ELISA method. For seroprevalence studies IgG is an accepted and reliable tool and indicates past exposure to a pathogen and determines the overall prevalence of a disease within a population.

### Sampling Strategy

Samples were selected using a stratified random sampling approach to ensure adequate representation across different age groups (0-18, 19-30, 31-45, 46-60, and >60 years) and both sexes. The samples were obtained from patients who had undergone routine laboratory investigations during their hospital visit. No specific clinical selection criteria were applied; however, patient demographics, including age, gender, and comorbidities, were retrieved from the electronic medical records (EMR) system.

### Inclusion Criteria

Serum samples from individuals across all age groups and both sexes.

Patients attending the hospital for routine or diagnostic investigations.

### Exclusion Criteria

Hemolysed or insufficient serum samples.

Samples from patients with known immunodeficiency or on immunosuppressive therapy.

### hMPV PCR Trends Study

Retrospective data from RT-PCR tests conducted at AIG Hospitals during the years 2023 and 2024 were analyzed. A total of 1,276 respiratory samples were tested for human metapneumovirus during this period (619 samples in 2023 and 657 samples in 2024).

### Testing Strategy

Respiratory virus testing, including hMPV, was performed as part of a syndromic diagnostic panel for patients presenting with respiratory symptoms. Testing was primarily conducted for patients with respiratory symptoms and those requiring hospitalization, Patient demographic data, including age, gender, comorbidities, co-infections, and hospitalization outcomes, were collected from the EMR system. Samples were from different age groups (0-18, 19-30, 31-45, 46-60, and >60 years) and both sexes representing a cross-section of the population.

### Inclusion Criteria

Patients with respiratory symptoms who underwent multiplex respiratory virus PCR testing.

Respiratory samples (nasopharyngeal swabs, sputum, bronchoalveolar lavage [BAL], or endotracheal aspirates) submitted to the laboratory during 2023 and 2024.

### Exclusion Criteria

Samples with inadequate clinical data.

Poor-quality samples or those with insufficient volume for testing.

### Ethical Statement

Written informed consent was obtained from all study participants prior to enrolment. The study was approved by the Institutional Ethics Committee (approval number: AIG/IEC-Post BH&R66/01.2025-01).

### Laboratory Methods

RT-PCR Assay:

Respiratory samples were tested using one of the following methods:

Biofire Pneumonia Panel (Filmarray Multiplex PCR assay, Biomerieux) for sputum, BAL, or ET samples. Samples were transported at 2-8°C and tested within 4 hours of receipt.

Respiratory Combined Panel (Multiplex PCR by TRUPCR, 3B BlackBio Biotech India Ltd.) for nasopharyngeal swabs. Samples were collected in 3 mL viral transport medium (VTM), transported at 2-8°C, and tested within 24 hours. Nucleic acid extraction was performed using the TRUPCR Total Nucleic Acid Extraction Kit, and PCR was conducted according to the manufacturer’s instructions. Following Viruses were included in both the panelsInfluenza A, B&C,,Respiratory Syncytial Virus (RSV), Adenovirus, Parainfluenza viruses,Human Metapneumovirus (hMPV), Human Rhinovirus/Enterovirus,Coronaviruses ,Boca Virus, Parecho Virus, MERS-Cov ELISA Test:

Serum samples were separated and stored at 2-8°C. Testing was performed within 48 hours using the hMPV-IgG ELISA kit (Cat. No: SLY3967Hu, SUNLONG Biotech Co. Ltd.). This is an indirect ELISA test. The assay was performed according to the manufacturer’s instructions. The mean optical density (OD) value and standard deviation (SD) of negative control serum were calculated. The cut-off value was determined using the formula:

### Cut-off Value = Mean OD of negative control + 3SD

A sample was considered positive if its OD value exceeded the cut-off.

### Statistical Analysis

The relationship between gender (male/female) and outcome (positive/negative) across different age groups was analyzed using the chi-square test for independence in GraphPad Prism 8.0. Data were organized into contingency tables, and a p-value ≤ 0.05 was considered statistically significant. Graphical representations, including bar graphs and contingency plots, were generated to visualize the results.

## RESULTS

### Seroprevalence of hMPV IgG Antibodies

A total of 820 serum samples were tested for the presence of hMPV IgG antibodies, comprising 411 samples from males and 409 from females. The overall seropositivity was 53.4% (438/820). Notably, individuals aged over 60 years exhibited a higher seropositivity rate, with a significant female predominance (p=0.001). In the 19–30 years age group, females demonstrated a significantly higher seropositivity rate compared to males (p=0.003). Conversely, in the 31–45 years age group, males showed a higher seropositivity rate (p=0.001). Detailed demographic data and antibody test results are presented in Table 1.

**Table: 1.** Demographic Details and HMPV IgG Antibody Positivity.

| Age groups | Total Positive | Male Positive(Males Positive/Total Males tested) | Female Positive(Females Positive/Total Females tested) |
| --- | --- | --- | --- |
| 0-18 yr | 88/165 (53.3%) | 42/82 (51.2%) | 46/83 (55.4%) |
| 19-30 yr | 56/165 (33.9%) | 19/82 (23.2%) | 37/83 (44.6%) |
| 31-45 yr | 86/164 (52.4%) | 54/83 (65.1%) | 32/81 (39.5%) |
| 46-60 yr | 81/162 (50%) | 39/81 (48.1%) | 42/81 (51.9%) |
| >60 yr | 127/164 (77.4%) | 61/83 (73.5%) | 66/81(81.5%) |
| **Total** | **438/820 (53.4%)** | **215/411 (52.3%)** | **223/409 (54.5%)** |

Where N= Total tested

### hMPV PCR Positivity

In year 2023, 619 samples were tested for respiratory viruses using PCR, with an hMPV positivity rate of 7.8% (48/619). In 2024, among 657 samples tested, the positivity rate decreased to 3.8% (25/657).

Demographic details and hMPV PCR test results for both years are given in Table 2 (2023 & 2024).

**Table:2.**

| Table:2 | hMPV PCR Data-2023 |  |  | hMPV PCR Data-2024 |  |  |
| --- | --- | --- | --- | --- | --- | --- |
| Age groups | Total Positive | Male Positive (Males Positive/Total Males tested) | Female Positive (Females Positive/Total Females tested) | Total Positive | Male Positive (Males Positive/Total Males tested) | Female Positive (Females Positive/Total Females tested) |
| 0-18 yr | 0/9 | 0/5 | 0/4 | 0/21 | 0/10 | 0/11 |
| 19-30 yr | 4/32 (12.5%) | 4/23 (17.4%) | 0/9 | 0/30 | 0/17 | 0/13 |
| 31-45 yr | 4/70 (5.7%) | 4/46 (8.7%) | 0/24 | 2/81 (2.5%) | 2/39 (5.1%) | 0/42 |
| 46-60 yr | 7/148 (4.7%) | 4/101 (4%) | 3/47 (6.4%) | 6/144 (4.2%) | 2/86 (2.3%) | 4/58 (6.9%) |
| >60 yr | 33/360 (9.2%) | 20/260 (7.7%) | 13/100 (13%) | 17/381 (4.5%) | 15/253 (5.9%) | 2/128 (1.6%) |
| Total | 48/619 (7.8%) | 32/435 (7.4%) | 16/184 (8.7%) | 25/657 (3.8%) | 19/405 (4.7%) | 6/252 (2.4%) |

Where N= Total tested

### Seasonal Variation

Monthly hMPV PCR positivity rates for 2023 and 2024 are detailed in Tables 3, and illustrated in Figures 7. A distinct seasonal pattern was observed, with infection peaks occurring in February–March and October.

**Table 3:** Month wise hMPV PCR percent Positivity-2023 & 2024.

| **Month**<br>**Wise** | **hMPV PCR percent Positivity-2023** |  | **hMPV PCR percent Positivity-2024** |  |
| --- | --- | --- | --- | --- |
|  | **No. of Positive for**<br>**HMPV/Total**<br>**samples tested)** | **HMPV**<br>**Percentage (%)** | **No. of Positive for**<br>**HMPV/Total**<br>**samples tested)** | **HMPV**<br>**Percentage (%)** |
| **Jan** | 2/68 | 2.94 | 1/49 | 2.04 |
| **Feb** | 7/81 | 8.64 | 3/77 | 3.89 |
| **Mar** | 7/60 | 11.6 | 2/85 | 2.35 |
| **Apr** | 0/34 | 0 | 1/61 | 1.63 |
| **May** | 0/22 | 0 | 0/35 | 0 |
| **Jun** | 0/25 | 0 | 0/29 | 0 |
| **Jul** | 1/30 | 3.33 | 0/45 | 0 |
| **Aug** | 5/51 | 9.8 | 5/79 | 6.32 |
| **Sep** | 9/81 | 11.11 | 5/84 | 5.95 |
| **Oct** | 13/84 | 15.47 | 6/46 | 13.04 |
| **Nov** | 2/44 | 4.54 | 1/35 | 2.85 |
| **Dec** | 2/39 | 5.12 | 1/32 | 3.12 |
| **Total** | **48/619** | **7.8** | **25/657** | **3.8** |

**Fig-1.**
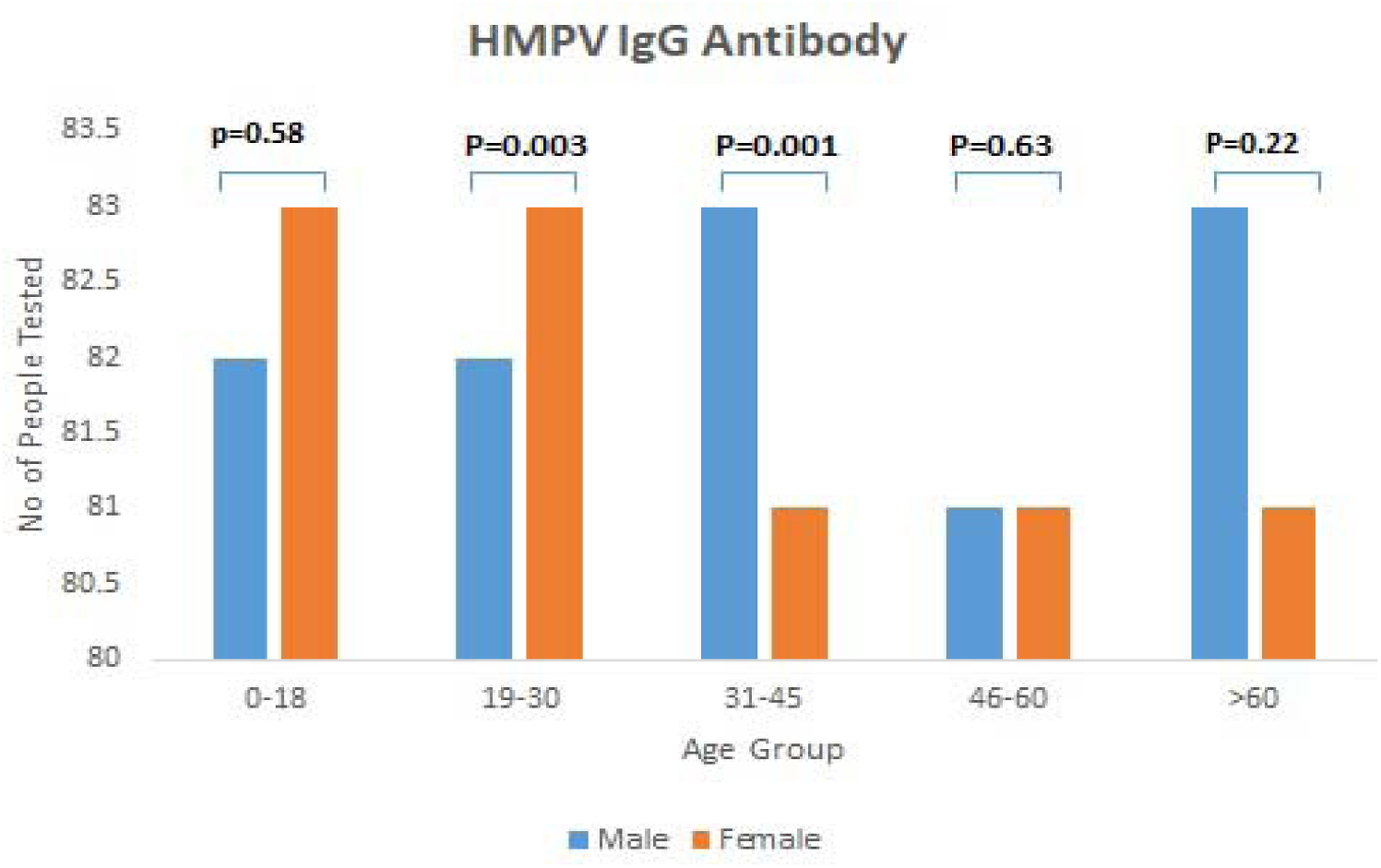
SEX DISTRIBUTION OF HMPV IgG ANTIBODY.

**Fig-2.**
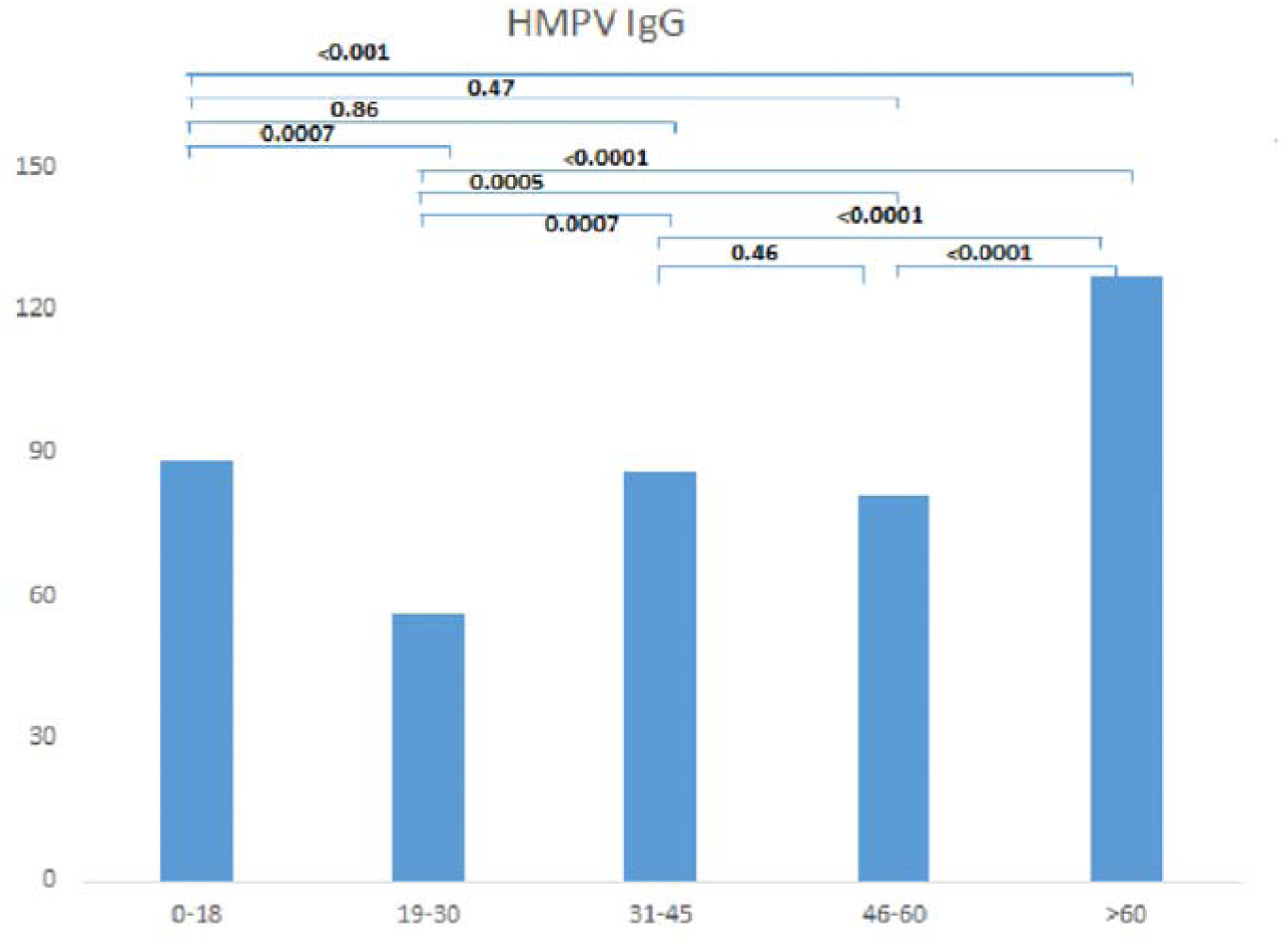
AGE DISTRIBUTION OF HMPV IgG ANTIBODY.

**Fig-3.**
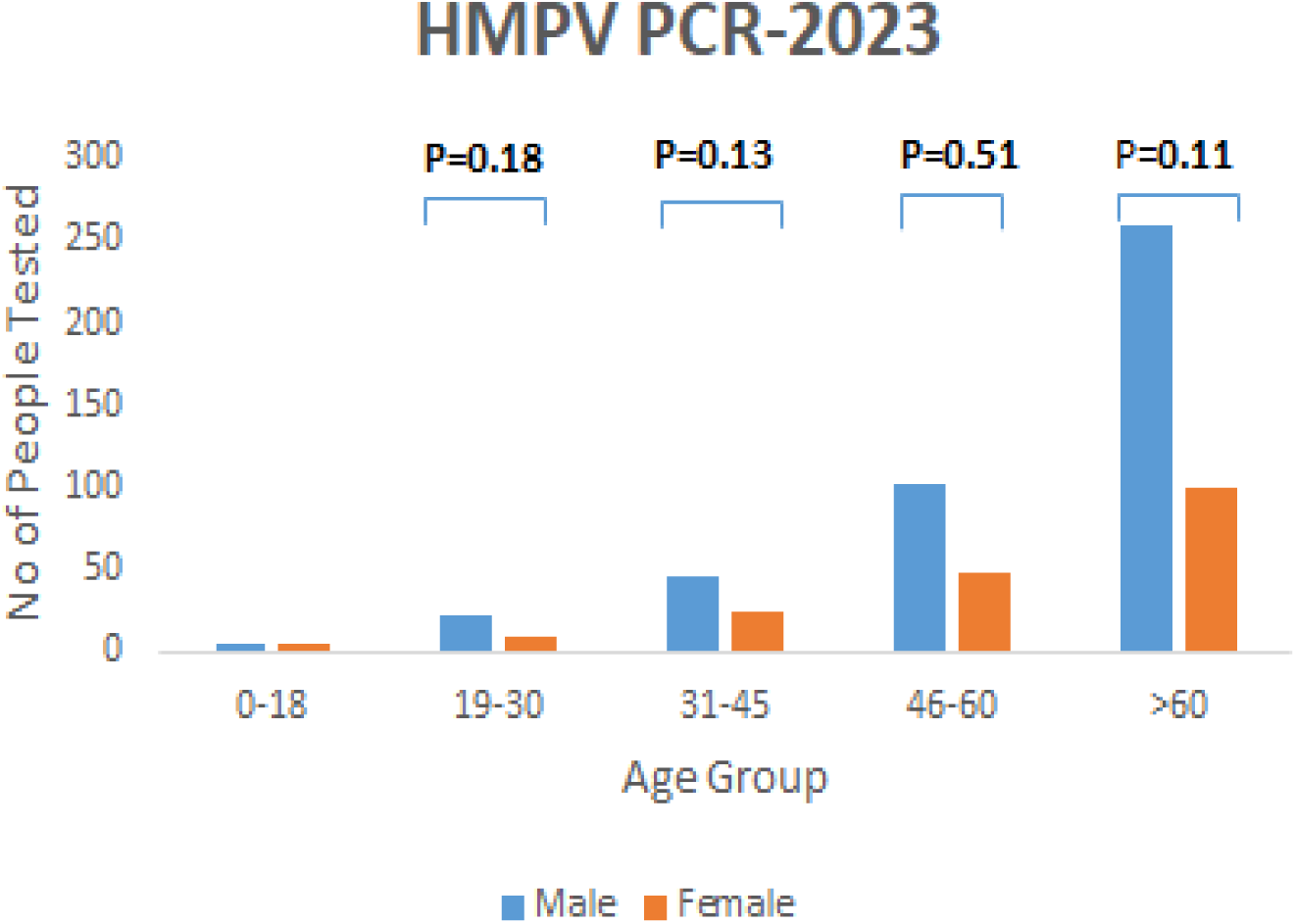
SEX DISTRIBUTION OF RT PCR 2023.

**Fig-4.**
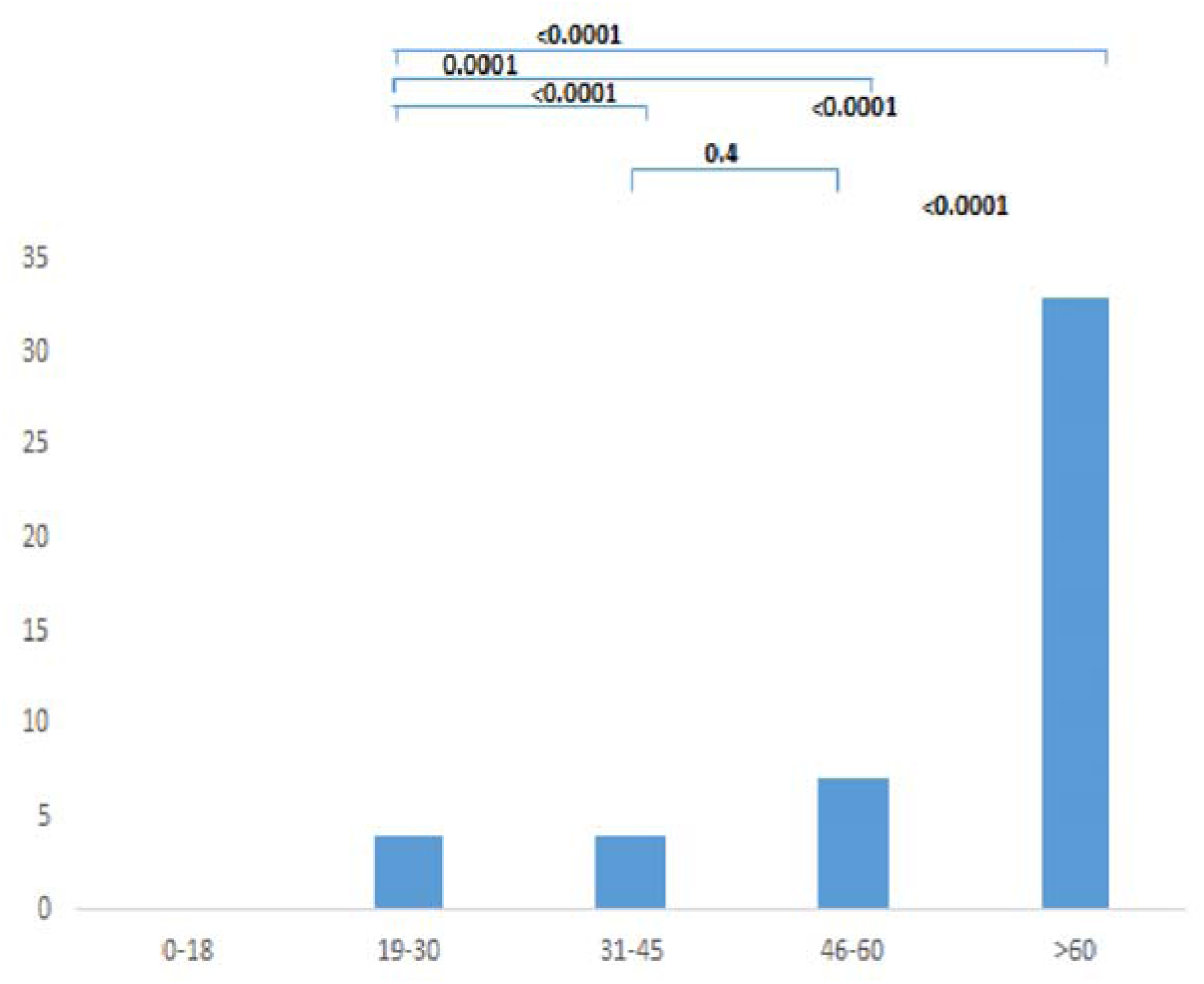
AGE DISTRIBUTION OF RT-PCR 2023.

**Fig-5.**
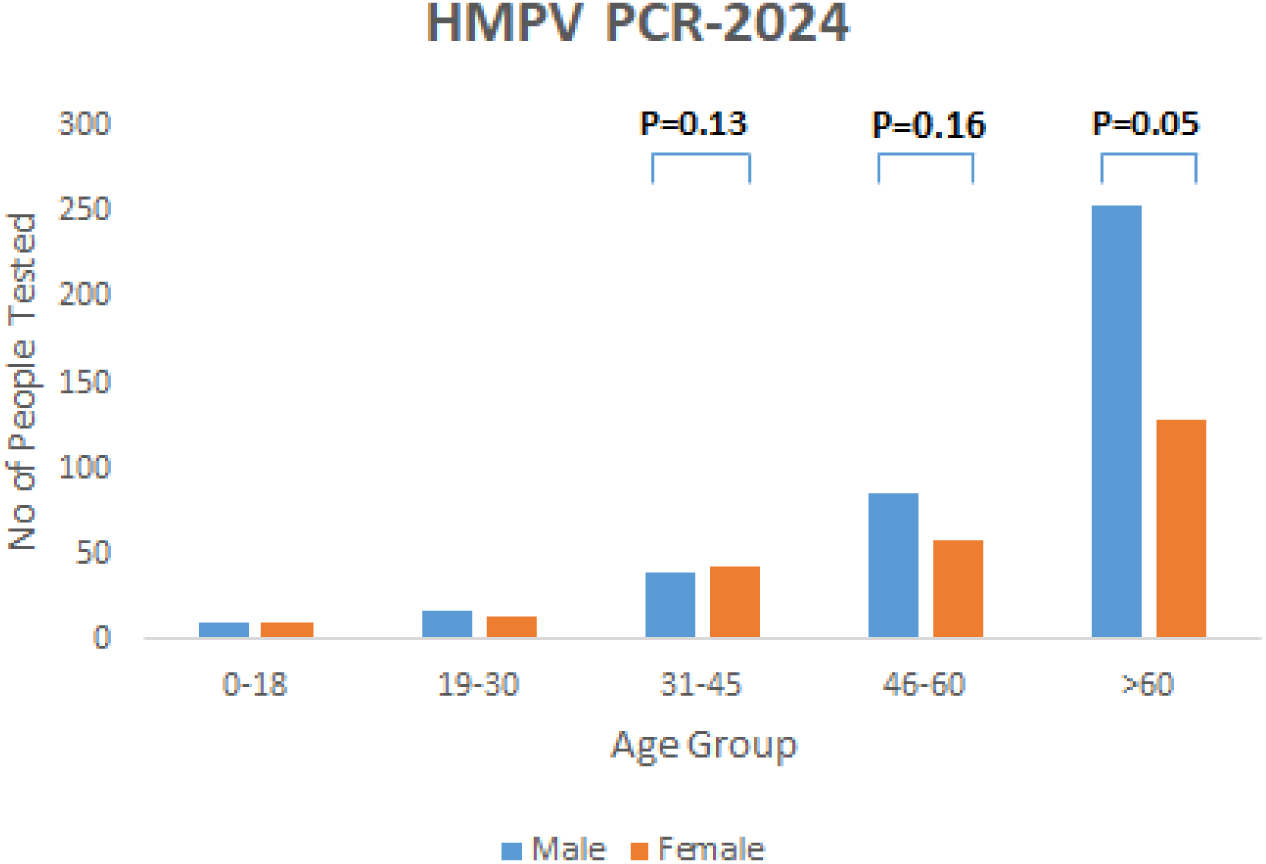
SEX DISTRIBUTION OF RT-PCR 2024.

**Fig-6.**
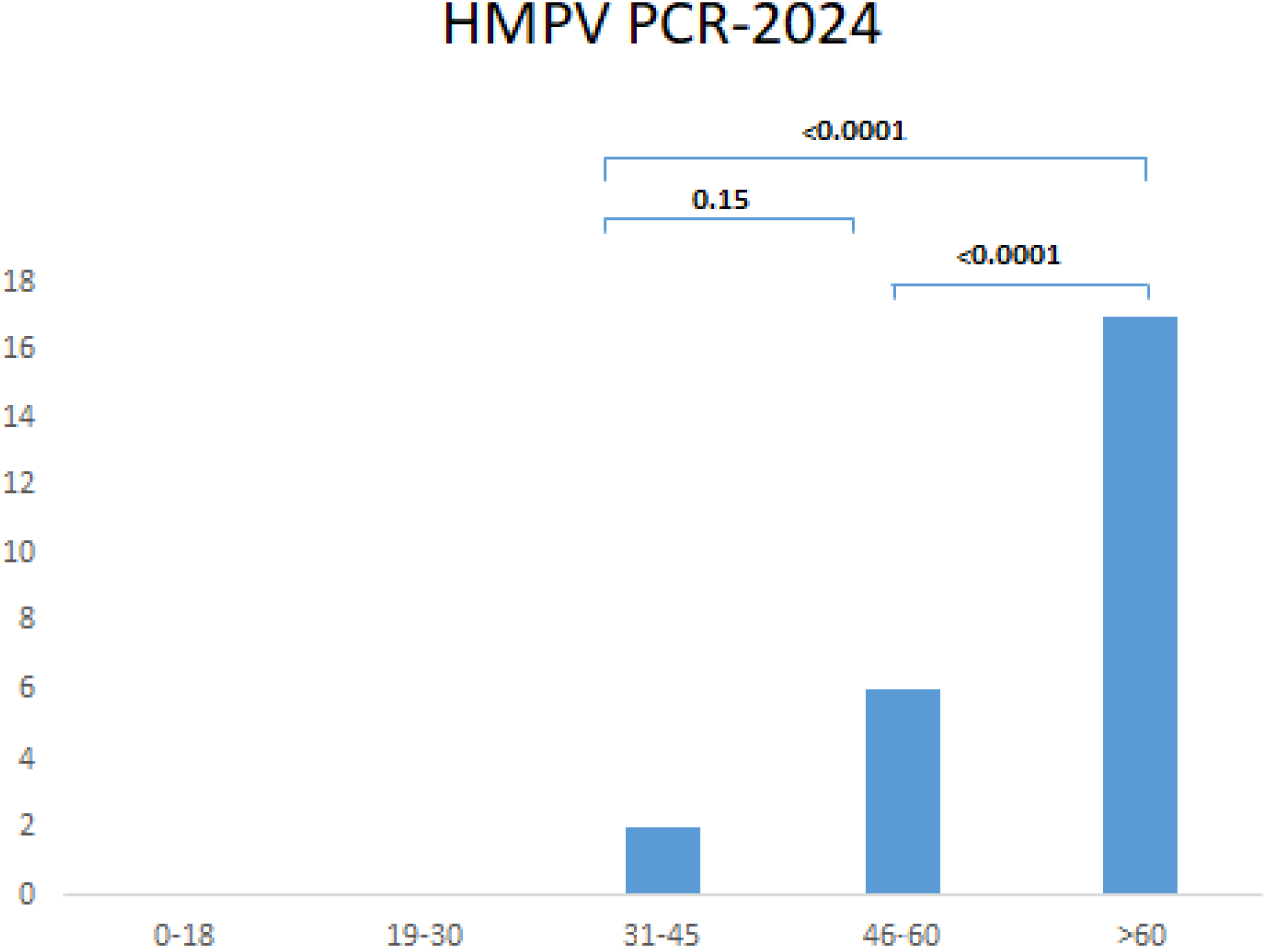
AGE DISTRIBUTION OF RT-PCR 2024.

**Fig-7.**
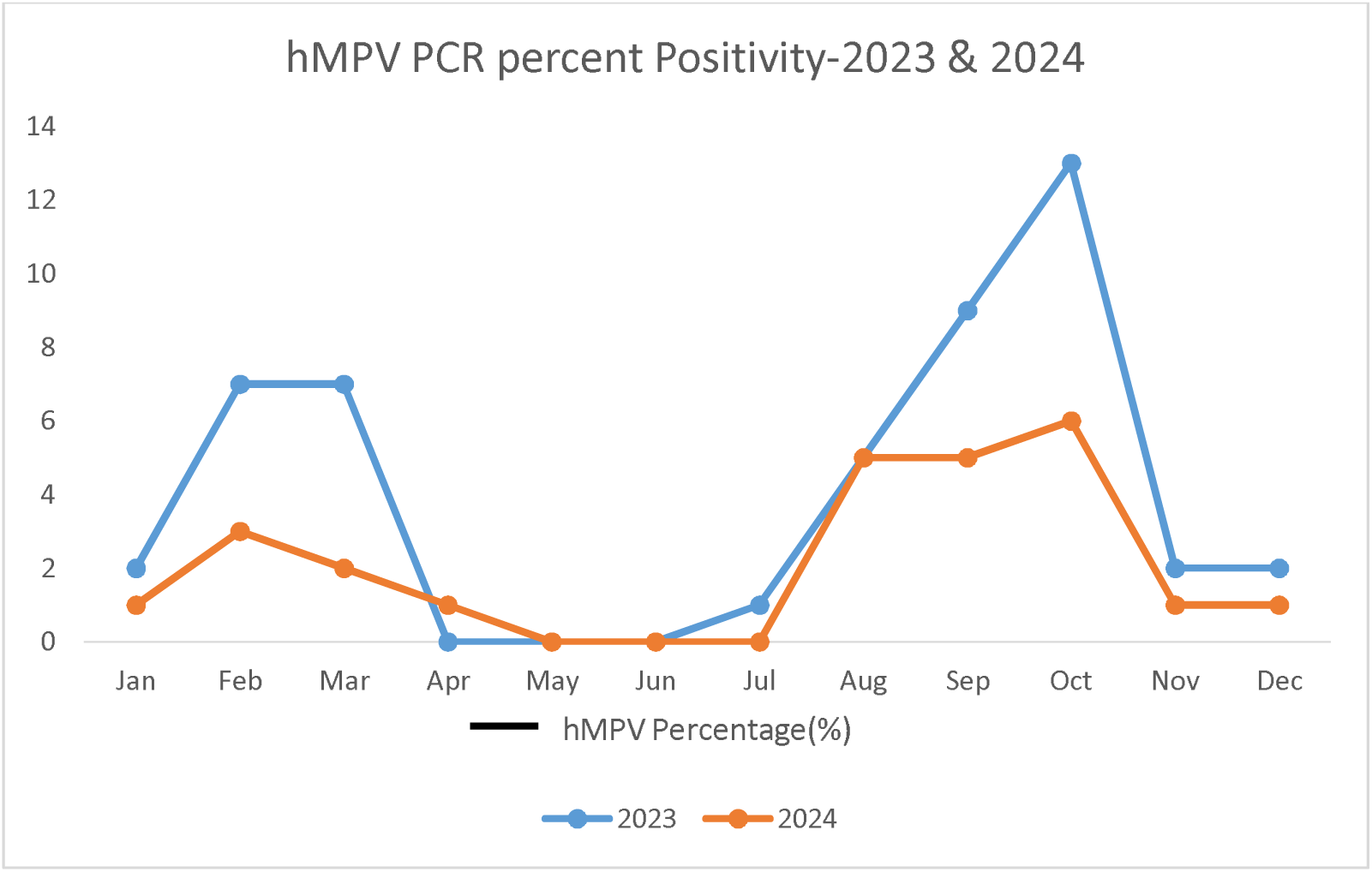

**Fig-8.**
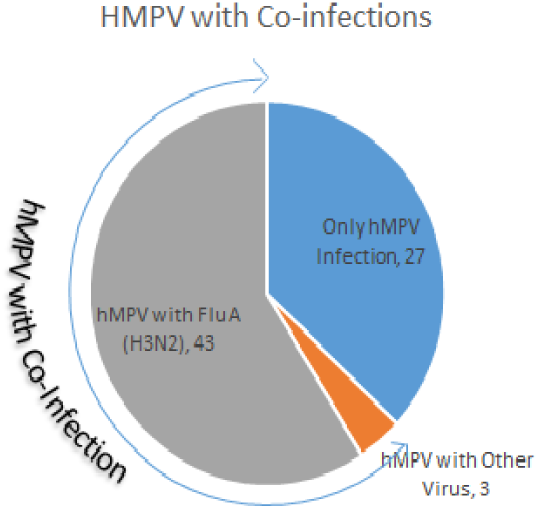
DISTRIBUTION WITH CO-INFECTIONS.

**Fig-9.**
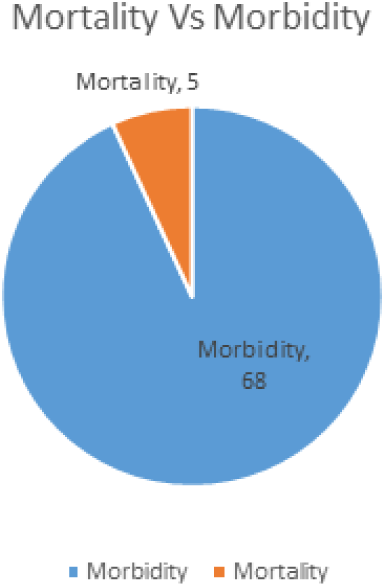
DITRIBUTION WITH MORBIDITY AND MORTALITY.

### Demographics and Clinical Outcomes of hMPV PCR-Positive Patients

Combining data from 2023 and 2024, a total of 73 patients (51 males and 22 females) tested positive for hMPV by PCR. The age distribution and demographic details are summarized in Table 4. The highest positivity rate, 6.7% (50/741), was observed in patients aged over 60 years.

**Table: 4.**
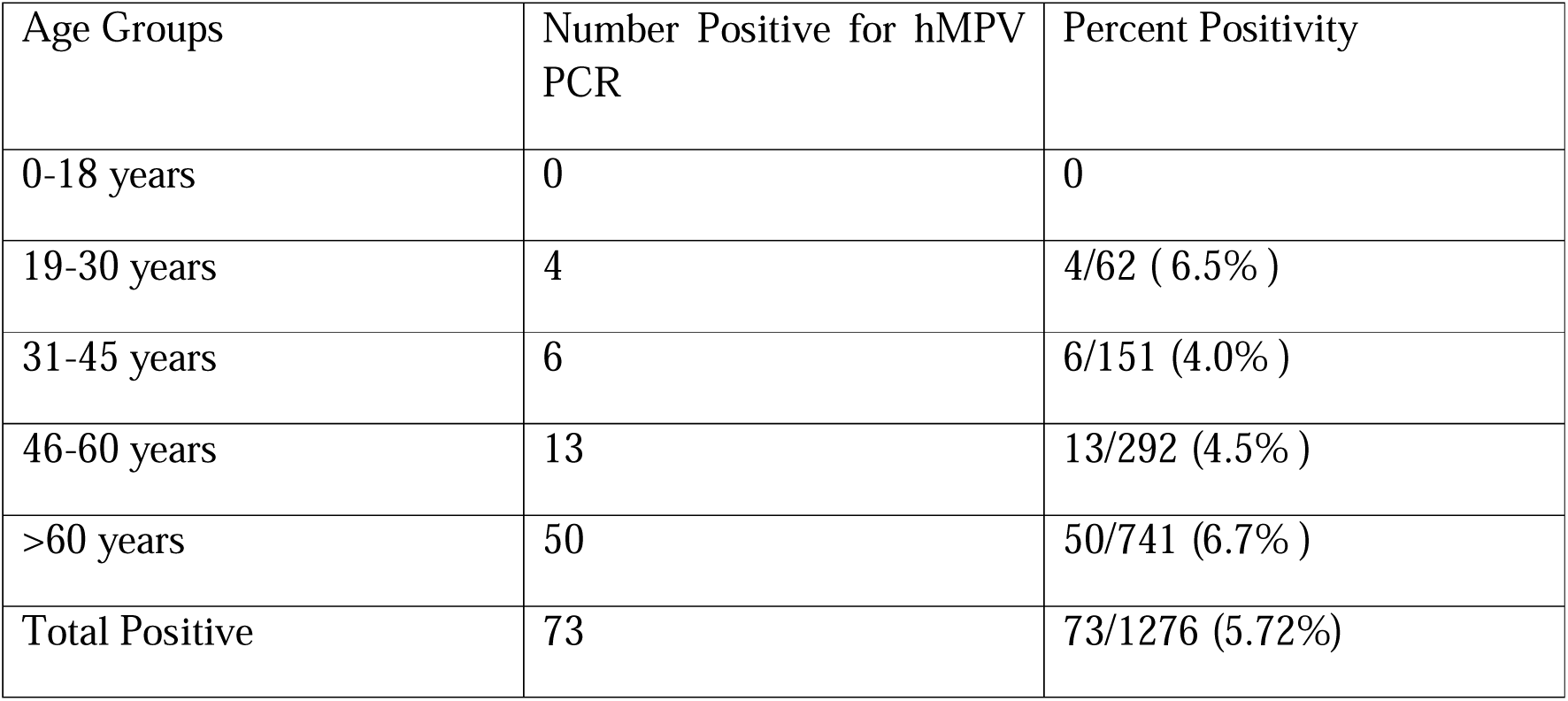
Age wise distribution of 73 Patients who tested Positive for hMPV by PCR in the years 2023 & 2024(Combined)

| Age Groups | Number Positive for hMPV PCR | Percent Positivity |
| --- | --- | --- |
| 0-18 years | 0 | 0 |
| 19-30 years | 4 | 4/62 ( 6.5% ) |
| 31-45 years | 6 | 6/151 (4.0% ) |
| 46-60 years | 13 | 13/292 (4.5% ) |
| >60 years | 50 | 50/741 (6.7% ) |
| Total Positive | 73 | 73/1276 (5.72%) |

Admission and Outcome status of positive patients is given in Table 5.

**Table :5.** Admission and Outcome status of 73 Patients who tested Positive for hMPV by PCR in the years 2023 & 2024(Combined)

| Admission status | In-Patient | Out Patient |
| --- | --- | --- |
| Total Number | 68 /73 (93.2% ) | 5/73 (6.8% ) |
| Deaths | 5/68 (7.4% ) | - |
| Discharges | 63/68 ( 92.6% ) | - |

Clinical outcomes indicated that 93.2% of hMPV-positive patients required hospitalization, with a mortality rate of 7.4% among these cases. Table 5. 92.6% (63/68) got discharged and were doing well on follow up.

### Co-infections and Comorbidities

Among the 73 hMPV PCR-positive patients, 63% (46/73) had comorbidities and other viral co-infections, with Influenza A (H3N2) being the most common co-infecting virus (93.5%). The remaining 37% (27/73) were infected solely with hMPV, among whom 18.5% (5/27) succumbed to the infection.

Of the 5 patients who died, the median age was 69 years (range: 56-78). All had underlying comorbidities, including chronic Liver disease (CLD) (n=1), diabetes mellitus (n=2), and CKD (n=2). The primary causes of death were respiratory failure (n=3), sepsis (n=1), and cardiac arrest (n=1).

## DISCUSSION

A Cross-sectional seroprevalence study conducted in the month of January 2025 from 25^th^ to 31^st^ showed that 53.4% of the study population had detectable IgG antibodies to hMPV indicating exposure to the Virus.

Our study indicates that among the 820 samples tested 53.3% (88/165) of subjects aged 0-18 years, 33.9% (19/30) aged 19-30 years, 52.4% (86/164) aged 31-45 years,50% (81/162) aged 46-60 years and 77.4% (127/164) aged >60 years were seropositive for hMPV. There was only a marginal difference in seropositivity in males (52.3%) and Females (54.5%). An Indian Study by Banerjee et al, (^4)^ from Delhi in the Year 2011 reported an overall percent positivity of 79% (79/100) whereas our study shows 53.4% (438/820) Positivity. Guilan Lu et al ^(5)^ in 2011 reported 100% Positivity in subjects 20 years and above. The lower seroprevalence in our study may reflect regional variations, differences in study periods, or a decline in hMPV circulation over time.

Prevalence of hMPV may differ between geographical locations. There are no seroepidemiological studies from Southern India. Our results vary from the one report published from India in 2011 ^(4)^. Number of Paediatric patients attending our hospital is considerably lower than adults. Hence we did not have data for subjects below 1 year. There is a detectable low seroprevalence among the age group 19 -30 years. In the 19-30 years age group females had comparatively higher seroprevalence (44.6 %) than males (23.2%). Whereas in the 31-45 years age group it was opposite, higher percentage of males (65.1%) had antibodies than females (39.5%). The reason for these differences is unknown but may be due to the varying levels of exposure and immunity while IgG ELISA is a standard method to assess seroprevalence, it primarily reflects humoral immunity. Given that HMPV is thought to elicit a significant cell-mediated immune response, the absence of detectable IgG may not necessarily indicate a lack of previous exposure. Jennifer E.Schuster et al have shown that antibody responses to hMPV can wane over time, leading to potential reinfections .

There is seasonal variation in hMPV percent positivity. From April to July the percent positivity of hMPV is very low in both years with no cases detected in May and June in both years (Table 3 and Figures 7)

From August the percent positivity increases gradually with a peak in October (late Monsoon) tapering off from November to January and gradually increasing again showing a second peak from February to March (Late winter). Mridu Anand et al, ^(3)^ reported a similar bimodal peak in incidence of respiratory viruses from Hyderabad during their study period (2017–2019)

In year 2023 of the total 619 tests done 70.3% (435/619) were males and 29.7% (184/619) were females. However overall percent positivity was slightly higher in females 8.7% (16/184) compared to males 7.4% (32/435). Table 2 In the year 2024 the number of males who got tested showed a slight decrease 61.6% (405/657) and there was an increase in the number of females who got tested 38.4% (252/657). The percent positivity has come down significantly in both males 4.7% (19/405) and females 2.4% (6/252) Table 2.

In both years, hMPV was not detected in females up to the age of 45 years. However, antibodies were detected when tested in January 2025 which indicates asymptomatic infections and circulation of the virus.

Over the period of two years it is observed that the PCR positivity has come down by 50%, from 7.8% in 2023 to 3.8% in 2024. Nandhini Et al ^(2)^ in 2016 reported similar findings from Puducherry, India during their study period from 2011 – 2013. They reported a percent Positivity of 15% in 2011, 5% in 2012 and 3% in 2013.

In our study, analysis of hMPV PCR Positive patients showed that 93.2% of them were hospitalised, of which 7.4% (Table.5) expired. P. Loubet et al in a study from France reported a mortality of 4% in hospitalised patients whereas Sang-Ho Choi et al reported an in-house mortality of 38.0%.

Of the 73 hMPV PCR Positive patients 46 (63%) had comorbidities and other virus co-infections of which Influenza A (H3N2) was the highest (93.5%) (Supplemental Table S1). 27 patients (37%) were infected with only hMPV out of which 5 (18.5%) had expired. Edward E. Walsh et al reported dual viral infections in 22.9% in hospitalised patients and 7% deaths in hMPV alone infected patients.

A total of 1276 patients were tested in years 2023 and 2024 for respiratory viruses by PCR. In 780 patients no respiratory virus was detected and mortality was 19.7% (154/780). 423 patients were hMPV negative but positive for other respiratory viruses and the mortality was 13.2% (56/423). In 73 patients hMPV was detected and mortality was 6.8% (5/73). Viral co-infections, where patients have more than one virus at the same time, are common. The impact of these co-infections on the severity of illness is still being debated. However, Nandhini et al in their study did not report any fatalities in their study.

The full cohort of 1276 PCR tested patients were not analysed for clinical details (like symptoms, severity, Outcomes and comorbidities). However, from the above data it appears that like other respiratory viruses hMPV infections may be a co-factor in increased morbidity and mortality in vulnerable population.

### Limitations

This single-center study’s finding may not be generalizable to broader populations. The qualitative assessment of antibodies without quantification or evaluation of neutralizing capacity limits insights into protective immunity levels. The absence of hMPV genotyping restricts understanding of circulating lineages and their potential impact on disease severity. Furthermore, the limited representation of patients aged 0–5 years hinders assessment of hMPV burden in this vulnerable age group.

### Strengths

Despite these limitations, Our study’s large sample size and seroprevalence studies offer valuable insights into hMPV exposure and infection patterns, directing future immunological research and public health strategies like Outbreak preparedness

## Conclusion

Our study indicates that 53.4% of the population had detectable hMPV IgG antibodies, suggesting significant exposure to the virus. The lower seroprevalence compared to earlier studies may reflect reduced viral circulation and waning immunity over time. Higher hMPV

PCR and antibody positivity rates in individuals over 60 years, especially those with comorbidities, underscore the need for targeted interventions. The observed seasonal peaks in hMPV infections during February–March and October highlight the importance of continuous surveillance. Given the significant morbidity and mortality associated with hMPV, particularly among high-risk groups, the development and implementation of an effective vaccine should be prioritized.

## Data Availability

All data produced in the present study are available upon reasonable request to the authors.

## ACKNOWLEDGMENTS

The authors acknowledged the patients who participated in the study. The authors are also grateful to the institution from where we collected our data and authors/editors/publishers of all those articles, journals, and books from where the literature for this article has been reviewed and discussed. This study was supported by all the technical staff of Biochemistry and Microbiology departments of AIG Hospitals.

## Authors’ Contribution

Dr. Deepika and Dr. Sadhana Designed the study and analysed the data. And also prepared the Manuscript and Performed the Statistical analysis.

Dr.Ankit helped in preparing the manuscript

Dr. Nagamani , Dr. Sushma Collected the clinical data. Mr. Rajesh Performed the Tests

Dr. D Nageshwar Reddy gave Administrative support and made Critical analysis. Dr. Deepika, Dr. Sadhana, helped in final editing and approved the Manuscript.

